# Fast SARS-CoV-2 detection protocol based on RNA precipitation and RT-qPCR in nasopharyngeal swab samples

**DOI:** 10.1101/2020.04.26.20081307

**Authors:** COVID-19 Basque Inter-institutional Group (coBIG), coordinators and participants:, Xabier Guruceaga, Amanda Sierra, Daniel Marino, Izortze Santin, Jon Ander Nieto-Garai, Jose Ramón Bilbao, Maier Lorizate, Patricia Aspichueta, Adhara Gaminde-Blasco, Adrian Bozal-Leorri, Agustín Marín-Peña, Ainara Castellanos-Rubio, Ainhoa Iglesias-Ara, Aitor Rementeria, Ana Bernal-Chico, Andoni Ramirez-Garcia, Ane Olazagoitia-Garmendia, Asier Benito-Vicente, César Martín, Eider Bilbao Castellanos, Eneritz Rueda-Alaña, Fabio Cavaliere, Guiomar Perez de Nanclares, Igor Aurrekoetxea, Iraia Garcia-Santisteban, Irantzu Bernales, Itziar Gonzalez-Moro, Jan Tønnesen, Jimena Baleriola, Jon Vallejo-Rodríguez, Leire Aparicio-Fernandez, Leire Martin-Souto, Luis Manuel Mendoza, Maialen Areitio, Maialen Sebastian-delaCruz, Maren Ortiz-Zarragoitia, Mari Paz Serrano-Regal, Miren Basaras, Nora Fernandez-Jimenez, Olatz Pampliega, Santos Alonso Alegre, Susi Marcos, Teresa Fuertes-Mendizabal, Unai Galicia-Garcia, Uxue Perez-Cuesta, Verónica Torrano, Virginia Sierra-Torre, Xabier Buqué, Ugo Mayor

## Abstract

The SARS-CoV-2 pandemic has evolved far more aggressively in countries lacking a robust testing strategy to identify infected individuals. Given the global demand for fast and reliable diagnosis to determine the carrier individuals, a stock-out scenario for a number of essential reagents/kits used along the diagnostic process has been foreseen by many organizations. Having identified the RNA extraction step as one of the key bottlenecks, we tested several alternatives that avoid the use of commercial kits for this step. The analysis showed that 2-propanol precipitation of the viral RNA, followed by one-step RT-qPCR results in a sensitivity and specificity comparable to that provided currently by automatized systems such as the COBAS 6800 system. Therefore, this simple protocol allows SARS-CoV-2 testing independently of commercial kit providers in a time and cost-effective manner. It can be readily implemented in research and/or diagnostic laboratories worldwide, provided that patient confidentiality and researcher safety are ensured. Scaling up the testing capabilities of hospitals and research facilities will identify larger numbers of infected individuals to paint a clear picture of the COVID-19 prevalence, a pre-requisite for informed policy decision making.

## INTRODUCTION

The severe acute respiratory syndrome coronavirus 2 (SARS-CoV-2) is the causative agent of the Coronavirus Infectious Disease 2019 (COVID-19), which has been declared a pandemic in March 2020 by the World Health Organization (WHO). Data suggest that mortality appear to range between 0.5-8% depending on the availability of diagnostic testing and the capacity of the healthcare system (https://doi.org/10.1007/s11357-020-00186-0). SARS-CoV-2 has emerged as an extremely contagious virus. Transmission can occur through direct contact, droplet spray at short ranges, and by airborne transmission events at long-range distances by aerosol particles (https://doi.org/10.1146/an-nurev-virology-012420-022445).

Given the ability of the virus to transmit without direct person-to-person contact and the existence of asymptomatic (https://doi.org/10.3201/eid2607.200718), early release https://www.nature.com/articles/s41591-020-0869-5), or presymptomatic (http://dx.doi.org/10.15585/mmwr.mm6914e1) transmission events, control of SARS-CoV-2 spread poses serious challenges to healthcare systems. In response, on March 22^nd^ 2020, the WHO has recommended that all countries increase the number of tests carried out and has recognized that in order to overcome the shortages of testing reagents for diagnosis, new laboratory testing strategies must be developed (https://apps.who.int/iris/handle/10665/331509). Thus, extensive testing is one of the key anti-pandemic measures.

The gold-standard detection tools for SARS-CoV-2 rely on the detection of viral RNA by reverse transcription followed by quantitative polymerase chain reaction (RT-qPCR). Currently, most healthcare systems use highly automatized detection methods such as the COBAS 6800 system (Hoffmann-La Roche). These systems allow the processing of a large number of samples with a smaller workforce than manual protocols but rely on several reagents and kits, which are in a permanent danger of limited availability and even stock-out due to their increasing demand following the fast, global spread of SARS-CoV-2. As a response, and following WHO recommendations, alternative diagnostic protocols that do not rely on the use of kits suffering global shortages must be designed. RNA extraction has been identified as one of the key bottlenecks in the application of massive testing strategies due to the shortage of diagnostic kits. As a response, in this study several kit-free strategies for RNA extraction have been tested. Our analysis shows that RNA precipitation with 2-propanol followed by one-step RT-qPCR detection of SARS-CoV-2 using a combination of hydrolysis probes and intercalating dye technologies provides a complete and robust diagnostic protocol that could be scaled up for the implementation of massive testing in certified research labs. An updated version of the procedure can be found here: https://www.ehu.eus/es/web/umayor/covid-19-protocol-en.

## MATERIALS AND METHODS

### Ethical statement

The study was conducted at the University of the Basque Country (UPV/EHU, Leioa, Spain) in collaboration with the Cruces University Hospital after approval by the UPV/EHU Ethics Committee (project 2020/059, CEIAB M30/2020/074).

### Biological samples

Nasopharyngeal swab (NPS) samples collected in UTM (Universal Transport Medium) were diagnosed for SARS-CoV2 at the Microbiology Service of Cruces University Hospital (HUC, Barakaldo, Spain), which is part of the Basque Health system (Osakidetza). Before transport to our laboratories, all samples were anonymized using an alphanumeric code and inactivated with lysis buffer (COBAS omni lysis buffer, Ref. 06997538190; 1:1 in UTM), and then registered and transported by the Basque BioBank. For the initial set up of the protocol (Set #1), we received 60 NPS samples (30 positives and 30 negatives). In a second stage of blind validation, we used 105 additional NPS samples (Set #2), unaware of the diagnosis assessment made by HUC.

### Total RNA purification/extraction

Five RNA extraction methods were compared to ensure an enriched high RNA quantity/quality sample in the shortest time minimizing the exposure to toxic reagents (**Figure 1**): A,- DNAse treatment; B, RNA precipitation with 2-propanol; C, DNAse treatment followed by RNA precipitation with 2-propanol; D, phenol:chloroform:isoamylalcohol extraction; and E, phenol-based RNA extraction using Trizol-reagent (ThermoFisher, Waltham, MA, USA). RNA concentration and purity (260/280 ratio) was determined by UV spectrophotometry on a NanoDrop ND-1000.

**Figure 1.**
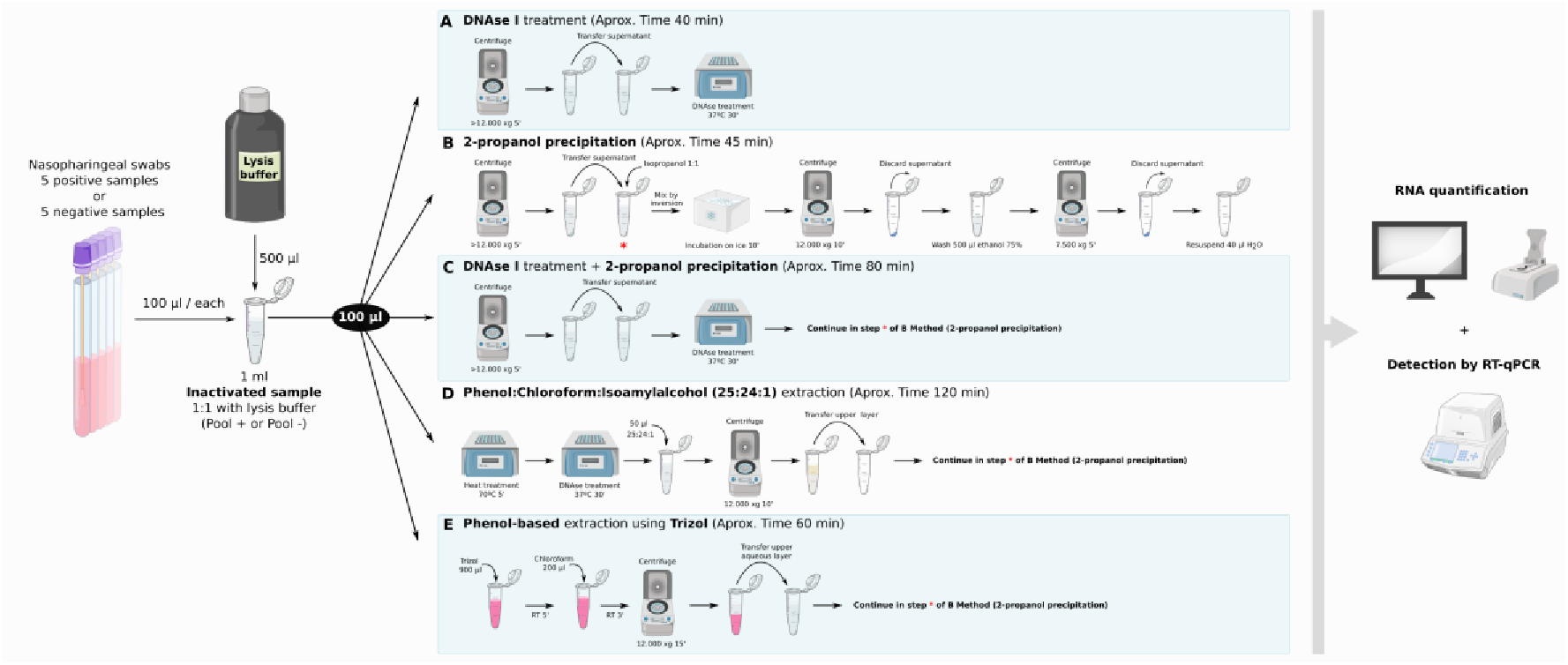
RNA extraction methods tested.

#### Detailed protocols are explained bellow

<u>(A) DNAse treatment:</u> An aliquot of 100 μl of inactivated sample was centrifuged for 5 min at 12,000 *g*. The supernatant was collected in a new 1.5 ml tube and was treated with 2 U of DNAse I (ThermoFisher) for 30 min at 37 °C following the manufacturer’s instructions.

<u>(B) 2-propanol precipitation:</u> An aliquot of 100 μl of inactivated sample was centrifuged and the supernatant collected as described in A. Then, the same volume (1:1) of pre-cold (−20 °C) 2-propanol (PanReac, Barcelona, Spain) was added, mixed by inversion and incubated for 10 min on ice. Afterwards, the sample was centrifuged for 10 min at 12,000 *g* and the supernatant was discarded. The pellet was washed in 500 μl of pre-cold (−20 °C) ethanol 75% (PanReac) and centrifuged for 5 min at 7,500 *g*. Finally, the pellet was air-dried for 10-20 min and resuspended in 40 μl of RNAse-free molecular grade water (PanReac).

<u>(C) 2-propanol precipitation after DNase I treatment:</u> This method corresponds to a combination of methods A and B. Thus, DNAse-treated samples as in A were further precipitated with 2-propanol as described in B.

<u>(D) Extraction with phenol:chloroform:isoamyl alcohol:</u> An aliquot of 100 μl of inactivated samples was heated at 70 °C for 5 min and treated with 2 U of DNAse I for 30 min at 37 °C. Then, 50 μl of phenol:chloroform:isoamylalcohol (25:24:1) (PanReac) were added and the mixture centrifuged for 10 min at 12,000 *g*. The aqueous upper layer was carefully recovered without disturbing the interphase and transferred to a new 1.5 ml tube. Finally, the samples were precipitated following the 2-propanol method described above.

<u>(E) Phenol-based extraction with Trizol reagent:</u> 900 μl of Trizol were added to an aliquot of 100 μl of inactivated sample and incubated 5 min at room temperature (RT). Then, 200 μl of chloroform (Sigma-Aldrich) were added, incubated for 3 min at RT, and centrifuged at 12,000 *g* for 15 min. The obtained aqueous layer was collected and transferred to a new 1.5 ml tube. Finally, the same volume of 2-propanol was added to continue with 2-propanol precipitation steps.

### SARS-CoV-2 detection by RT-qPCR with fluorogenic 5’ nuclease probes

Detection of SARS-CoV-2 in RNA samples from NPSs was performed with fluorogenic 5’ nuclease probes (hydrolysis probes), using the 2019-nCoV CDC qPCR Probe Assay (IDT, San José, CA, USA - CDC #225397445). This diagnostic panel is composed of a set of primers and probes designed and approved by the US Centers for Disease Control and Prevention (CDC, https://www.cdc.gov/) that target the viral gene *N* (encoding the nucleocapsid protein; probes N1, N2 and N3) and the human gene for the subunit 30 of the RNAseP *(RPP30)*, used to control for biological sample collection and nucleic acid extraction procedures. While the N1 and N2 probe sets were specific for SARS-CoV-2, the N3 probe set detected other coronaviruses, and was subsequently removed from the diagnostic panel by the CDC (https://www.cdc.gov/coronavirus/2019-ncov/lab/rt-pcr-panel-primer-probes.html; **Table 1**). As a negative control, RNA extracted from non-infected HeLa cells was included (30 ng/well). As a positive control, the commercial control plasmid 2019-nCoV_N_Positive Control (Ref. 10006625, IDT, Leuven, Belgium) was used. A non-template control (H_2_O) was also included. All samples and controls were run in duplicate.

**Table 1.**
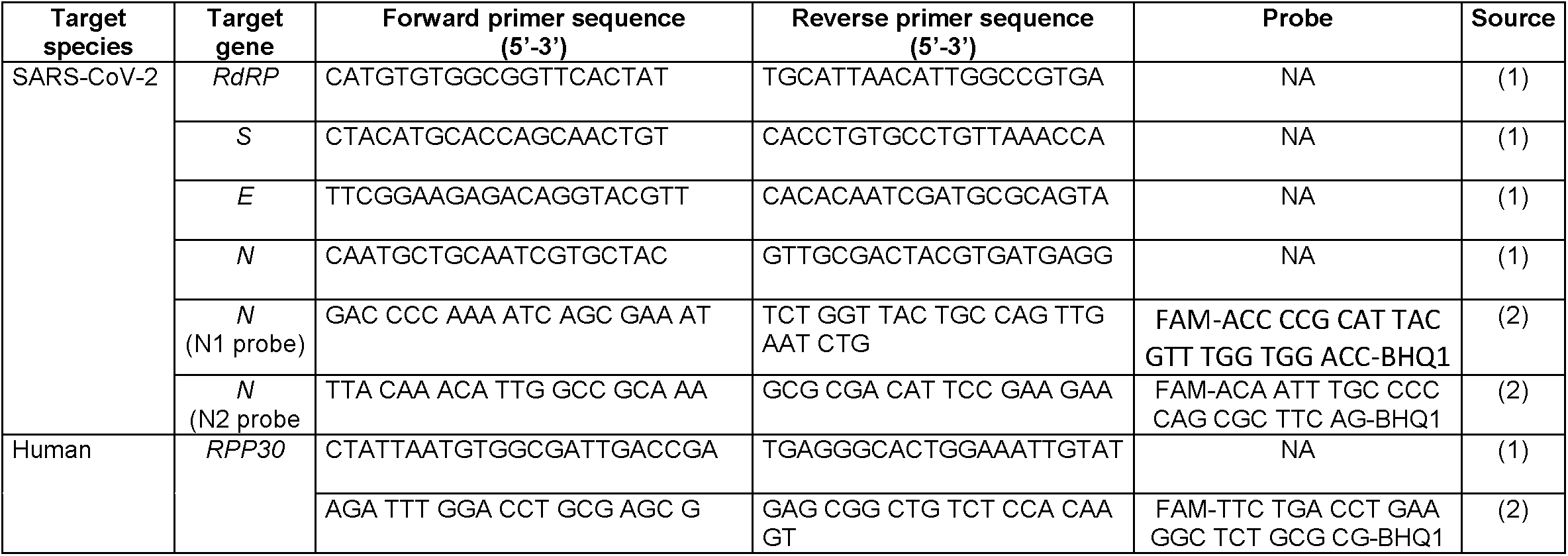
Sequence of primers and probes used. *The target species, target gene, sequence of forward and reverse primers for intercalating green agents, as well as the probe in fluorogenic 5’ nuclease assays are indicated. NA means not applicable. Source (1) indicates Won et al., 2020 (https://www.ncbi.nlm.nih.gov/pubmed/32156101) and Source (2) indicates CDC (https://www.cdc.gov/coronavirus/2019-ncov/lab/rt-pcr-panel-primer-probes.html)*.

RT-qPCR was performed using the NZYTech Speedy One-step RT-qPCR Probe Master Mix (NZYTech, Lisbon, Portugal), following the manufacturer’s instructions. Reactions were performed in 384-well plates in a final volume of 10 μl (2 μl RNA + 8 μl reaction mix) in a CFX384 Touch Real Time PCR Detection System (Bio-Rad, Hercules, CA, USA), using the following parameters: 20 min at 50 °C for reverse transcription of RNA; 3 min at 95 °C for polymerase activation and 40 cycles of amplification (95 °C 5 sec, 55 °C 50 sec). All samples and controls were run in duplicate. Annealing temperature was set to 55 °C following the recommendations of the CDC to allow viral detection in case of eventual mismatches due to mutations (https://www.fda.gov/media/134922/download).

### SARS-CoV-2 detection by RT-qPCR with an intercalating green dye

As an alternative SARS-CoV2 detection method for samples assigned as indeterminate, as defined below in the *Quality Control and Data Analysis* section, a method based on a intercalating green dye for detection of double-stranded DNA by RT-qPCR was used (NZYSpeedy One-step RT-qPCR Green Kit, NZYTech). We used three sets of primers previously described by Won et al. (https://www.ncbi.nlm.nih.gov/pubmed/32156101), that targeted viral genes: *N*, encoding for the nucleocapsid, *S* for the spike, and *RdRP* for the RNA-dependent RNA polymerase (**Table 1**). The primers for the *E* gene, which encodes for the envelope, were discarded because of their low specificity and hybridization in other viral targets based on BLAST (https://blast.ncbi.nlm.nih.gov/Blast.cgi). Primers for the human gene RNAseP subunit 30 *(RPP30)* were used as a control for biological sample collection and nucleic acid extraction. The reactions were run in duplicate in 384-well plates using 2 μl of RNA in a final reaction volume of 10 μl (QuantStudio 5 Real-Time PCR System;ThermoFisher, Waltham, MA, USA). The PCR program was 20 min at 50 °C for RNA reverse transcription; 3 min at 95 °C for polymerase activation and 40 cycles of amplification (95 °C 5 sec, 60 °C 50 sec), where 60 °C is the optimal annealing temperature for these primers. In addition, the analysis of the melting temperature curve (70–95 °C, in 0.2°C increments, 10 sec per step) was added at the end of the reaction as a control of amplification specificity. The negative and positive controls used for probe experiments were also included as controls for this assay.

### Quality Control (QC) and Data Analysis

#### 1. Probe methodology

An initial QC was performed before samples were analyzed for the presence of SARS-CoV-2 RNA. Non-template controls were required to be strictly negative for all assays, while the negative control (non-infected HeLa cells) had to be positive for the human *RPP30* gene (threshold cycle or Ct<35). Regarding the positive controls, the three serial dilutions of the standard curve had to be positive for both N1 and N2 viral probes and negative for the human *RPP30* endogenous control.

In anticipation of possible scenarios where scale up of analyses might be necessary, a semi-automated worksheet template that can estimate viral copy number and determine sample positivity according to the criteria outlined in **Table 2** was prepared (**Supplementary File 1**). In this script, raw experimental results are pasted into the template, standard curves for N1 and N2 are constructed from the positive control dilutions and the copy content of each viral gene is calculated for each clinical sample. Samples were considered positive for SARS-CoV-2 when N1 and/or N2 amplification could be detected and more than 4 copies per reaction were detected. In turn, samples were considered negative when neither N1 nor N2 amplification was detected. Finally, samples in which the amplification of *RPP30* was below the QC threshold (Ct>35), or showing non-concordant results between technical replicates, or presenting estimates of viral content below 4 copies (but Ct<40) were considered indeterminate for the presence of SARS-CoV-2 and their analysis was repeated with the intercalating green dye method.

Alongside the quantitative assignment of clinical samples into the positive, negative or indeterminate categories, an additional QC procedure was performed. The qPCR amplification curves for each sample were plotted from the relative fluorescence units (RFU) detected in each cycle, using an in-house R script (**Supplementary File 2**). The amplification curve of each indeterminate sample was examined and compared to positive and negative samples. Samples showing amplification curves that did not fit a sigmoidal function were considered negative for SARS-CoV-2 and were not repeated with the intercalating green dye experiment (**Supplementary Figure 1)**.

#### 2. Intercalating green dye methodology

In order to pass the initial QC, the non-template controls had to be strictly negative for all assays, while the negative control (non-infected HeLa cells) had to be positive only for the human *RPP30* gene (Ct<35). Regarding the positive control, all dilutions included in the standard curve had to show amplification for the *N* viral gene. Finally, all clinical samples had to display *RPP30* amplification with a Ct lower than 35, to ensure the quality of the sample and the nucleic acid extraction.

Following the criteria outlined in **Table 2**, samples were considered positive for SARS-CoV-2 when at least two of the three viral genes (*N*, *S* and/or *RdRP*) showed positive amplification (Ct<40) and negative when none of the viral genes amplified. Samples showing amplification of the *RPP30* endogenous control after the QC threshold (Ct>35), or showing non-concordant results between technical replicates, were considered indeterminate for the presence of SARS-CoV-2 and the need of a new sample collection will be informed.

**Table 2:** Criteria for the assignment of clinical samples into SARS-CoV-2 positive, negative or indeterminate categories. *The threshold cycle (Ct) and the number of copies of viral genes were used to determine whether samples were categorized as positive, negative or indeterminate. (-) indicates genes not evaluated because the sample does not meet minimal quality requirements*.

| Hydrolysis probe assay |  |  |
| --- | --- | --- |
|  | *RPP30* | **N1 and N2** |
| **Positive** | Ct < 35 | > 4 copies in N1 and/or N2 |
| **Negative** | Ct < 35 | < 4 copies and Ct > 40 in both |
| **Indeterminate** | Ct < 35 | < 4 copies but Ct < 40, in N1 and/or N2 |
|  | Ct < 35 | Discrepant technical replicates |
|  | Ct > 35 | - |
| Intercalating green dye assay |  |  |
|  | *RPP30* | **N, S and RdRP** |
| **Positive** | Ct < 35 | Ct < 40 in at least 2 genes |
| **Negative** | Ct < 35 | Ct > 40 in all 3 genes |
| **Indeterminate** | Ct < 35 | Samples not fulfilling pos. or neg. criteria |
|  | Ct < 35 | Discrepant technical replicates |
|  | Ct > 35 | - |

As an additional QC step, melting curve analysis of qPCR products was performed and wells with non-specific amplification products were eliminated from the assignment. The amplification curves of indeterminate samples were examined as described and those not fitting a sigmoidal function were considered negative for SARS-CoV-2.

### Statistical analysis

The results obtained with the test developed by the coBIG were compared with the results obtained by Osakidetza (considered herein as true positives and true negatives). Statistical analysis was performed using the DAG_Stat Excel file developed by Mackinnon (https://www.ncbi.nlm.nih.gov/pubmed/10758228). We calculated sensitivity (proportion of coBIG concordants positive over all Osakidetza positives samples), specificity (proportion of coBIG concordants negative over all Osakidetza negatives samples), and accuracy (as sum of concordant positives and negatives correctly identified by coBIG over total number of samples). Besides, we also determined the predictive value of positive (precision) or negative test, defined as the proportion of samples with a positive or a negative result that are correctly assigned, respectively. Finally, we calculated the false positive and negative rates, as positives or negatives incorrectly identified by coBIG over all positive or all negative Osakidetza samples, respectively. The Cohen’s Kappa index calculated measures the agreement between the two techniques excluding the possibility of the agreement occurring by chance. Calculations are explained in detail in **Supplementary Figure 2**.

## RESULTS

### Determination of the optimal kit-free total RNA extraction procedure

We first compared five RNA extraction methods (**Figure 1**) in two pools of positive and negative (5 samples each) NPSs from experimental Set #1. To select the most effective method for RNA extraction, three factors were taken into account: the time consumed, the RNA quantity/quality (Q/Q) ratio obtained (ratio 260/280 absorbance), and the usage of non-phenolic reagents to avoid toxicity (**Table 3**).

**Table 3.** Quantification of RNA isolated by the five extraction methodologies performed. *The method, the amount of RNA (ng/μl), and ratio 260/280 for both negative and positive pool samples are indicated. In addition, the approximate time to process 24 samples and whether RT-qPCR could successfully detect viral and human genes is also indicated. (A) DNAse I treatment; (B) 2-propanol precipitation; (C) 2-propanol precipitation + DNAse I treatment; (D) Phenol:Chloroform:Isoamylalcohol extraction. (E) Phenol-based extraction using Trizol; N/A: Undetermined values*.

| Method | Negative Pool |  | Positive Pool |  | Aprox. Time (min) | RT-qPCR qualifier |
| --- | --- | --- | --- | --- | --- | --- |
|  | RNA (ng/μl) | 260/280 | RNA (ng/μl) | 260/280 |  |  |
| A | N/A | N/A | N/A | N/A | 40 | Discordant |
| B | 13.8 | 1.53 | 15 | 1.76 | 45 | Good |
| C | 13.4 | 1.58 | 18.2 | 1.88 | 80 | Good |
| D | 6.1 | 1.51 | 14.1 | 1.93 | 120 | Good |
| E | 12.1 | 1.78 | 16.8 | 1.83 | 60 | Good |

From all the tested methods, direct precipitation of total nucleic acids (including RNA) using 2-propanol without DNAse I treatment (method B), was selected as the most appropriate as it was the second quickest (around 45 min), and showed good Q/Q values in the Nanodrop (**Table 3**) without using phenolic reagents. DNAse treatment (method A) was the fastest (around 40 min), but yielded little nucleic acids, as the Q/Q values of the RNA obtained were undetermined by NanoDrop and did not result in significant amplification of viral or human genes by RT-qPCR. Method C (a combination of A and B) required around 80 min and the Q/Q values obtained did not significantly improve those obtained with method B. Finally, phenol-based methods (methods D, E) showed good Q/Q values but they were disregarded because of the longer time needed and the toxicity of phenolic reagents used during the process.

### Comparison of SARS-CoV-2 detection protocols

After selecting direct 2-propanol precipitation (method B) as the optimal kit-free RNA extraction method we proceeded to compare SARS-CoV-2 by either one- or two-step RT-qPCR, or by using intercalating green dye primers or hydrolysis probes (**Figure 2, 3**). For this purpose, we first compared the detection threshold for the N gene using 1:10 serial dilutions of the positive control CoV-N down to 10^−5^. We found that both intercalating dye primers and hydrolysis probes had very similar amplification in the detection range (40,000–4 copies per well) (**Figure 2**).

**Figure 2.**
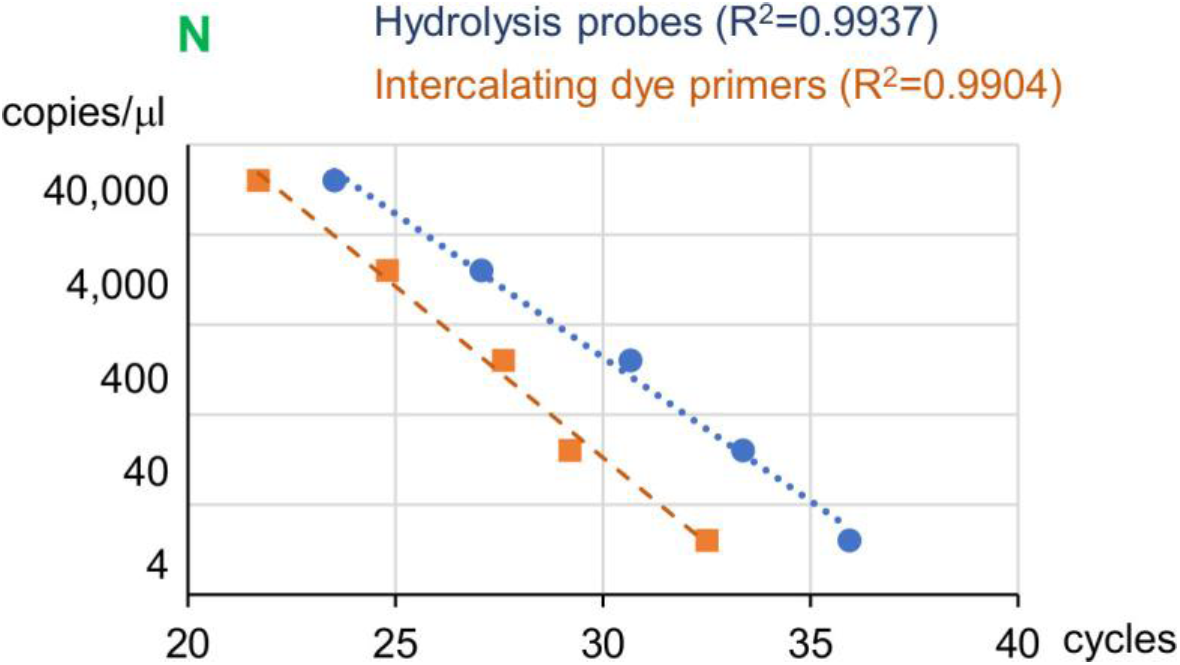
Standard calibration curve of hydrolysis probes and intercalating dye primers. *The amplification of the N gene was compared using the N1 hydrolysis probe (blue) and the N set of primers for intercalating gene (orange) assay using dilutions corresponding to 4–40,000 copies of N gene per PCR reaction. The determination coefficient (R*^2^*) for the linear adjustment is indicated*.

**Figure 3.**
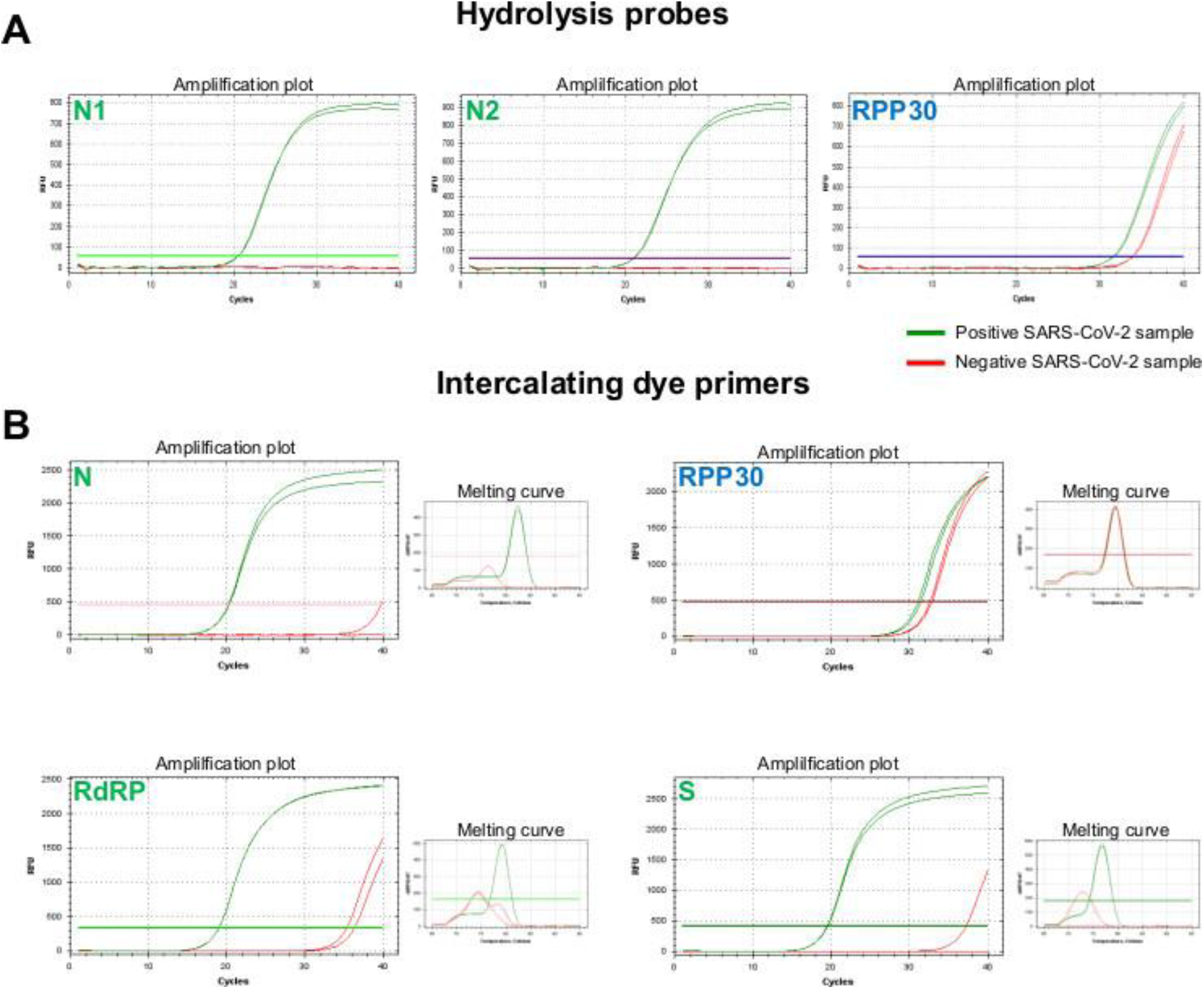
Comparison of amplification of positive and negative samples by hydrolysis probes and intercalating dye primers. ***A***, *hydrolysis probes amplification plots for viral genes (probes N1 and N2) and human gene (RPP30) for one positive SARS-CoV-2 sample (green) and one negative SARS-CoV-2 sample (red), in duplicate. Fluorescence thresholds are indicated by horizontal color lines*. ***B****, intercalating gene amplification plots and corresponding melting curves for viral genes (N, RdRP and S) and human gene (RPP30). Fluorescence thresholds are indicated by horizontal color lines*.

We then compared the detection of viral genes in some of the remaining NPS samples from Set #1 (n=24; 12 positive and 12 negative samples) using intercalating green dye primers and hydrolysis probes and found that both methods had a large degree of concordance (23/24 samples, 96% concordance; **Figure 3**). No significant effects of using one or two-step RT-qPCR reactions were found (data not shown). Finally, we used the whole remaining samples of Set #1 (n=46) to define our analysis criteria (**Table 2**). As the one-step RT-qPCR reaction lasted 1.5 h for hydrolysis probes and over 2 h for intercalating dye primers, we decided to base our protocol on the two hydrolysis probes for the *N* gene and to use the intercalating dye primers to double-check samples classified as indetermined with the hydrolysis probes assay (**Table 2**), as they would amplify three independent viral genes.

### Validation of the SARS-CoV-2 detection SOP

To validate our established standard operating protocol (SOP), we used sample Set #2, consisting of 105 already diagnosed and anonymized NPSs. To ensure a proper validation strategy our researchers were unaware of the diagnosis assessment made by Osakidetza. Therefore, RNA was extracted from the 105 NPS samples and SARS-CoV-2 was first detected by RT-qPCR using the fluorescent hydrolysis probes method. Following the criteria described in **Table 2**, the samples that did not fulfill the established criteria for an unequivocal diagnostic were further analyzed by testing three different viral genes (*N*, *S* and *RdRP*) using the intercalating dye RT-qPCR method. Overall, the combination of both detection methods permitted to establish final diagnostic for 94 of the 105 samples (89.5%) following the established diagnostic criteria, with 48 positives, 46 negatives and 11 indeterminate samples. We compared these results with the diagnostic results from Osakidetza, who diagnosed 47 positives, 44 negatives and 14 indeterminates. Thus, the coBIG protocol detailed here results in 91% sensitivity with 90% specificity in SARS-CoV-2 detection (**Table 4**). The Cohen’s kappa index is calculated as 0.81 indicating almost perfect agreement excluding chance.

**Table 4.**
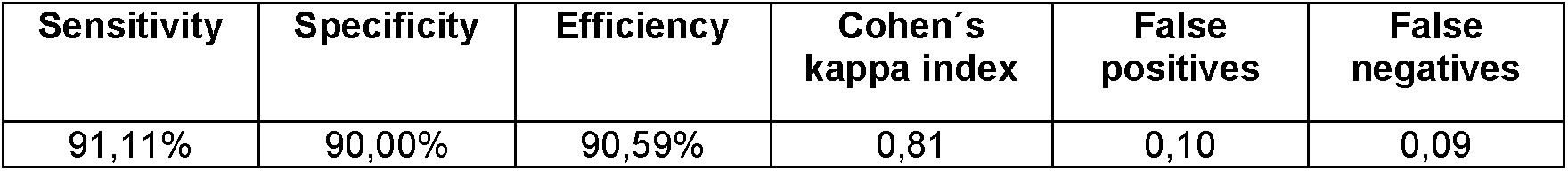
Summary of statistical analysis of 105 blind samples analyzed.

## DISCUSSION

In this manuscript, we have developed a method for in-house detection of SARS-CoV-2 based on 2-propanol precipitation of inactivated samples followed by one-step RT-qPCR using the CDC hydrolysis probes N1 and N2, and subsequent confirmation of indeterminate samples using intercalating dye primers for *N*, *S* and *RdRP*. We provide evidence that this method provides a 90% specificity and 91% sensibility compared to the diagnostic assessment made by the Basque Health Service, Osakidetza.

Massive SARS-CoV-2 testing is reclaimed by scientists worldwide to fight the current pandemic (https://www.nature.com/articles/d41586-020-00864-1). Without solid data about the prevalence of the COVID-19, epidemiological models will be flawed (https://www.nature.com/articles/d41586-020-01003-6), and the policies informed by them will be ineffective at the very best, and utterly dangerous for our health and economies at the worst. In fact, research labs worldwide are being repurposed to increase the testing capabilities for SARS-CoV-2 (https://www.nature.com/articles/d41586-020-00905-9) in initiatives similar to the one presented here. RT-qPCR is currently the gold-standard for detecting SARS-CoV-2 although novel genetic, protein, or serological methods are emerging (https://www.ncbi.nlm.nih.gov/pmc/articles/PMC7144809/). Novel genetic methods are based on CRISPR/Cas12 (Clustered Regularly Interspaced Short Palindromic Repeats) (https://www.nature.com/articles/s41587-020-0513-4) or LAMP (loop-mediated isothermal amplification) (in public repositories; reviewed in https://www.ncbi.nlm.nih.gov/pmc/articles/PMC7144809/). SARS-CoV-2 protein N detection also provides an alternative method for diagnostic but its sensibility is unclear (https://www.ncbi.nlm.nih.gov/pmc/articles/PMC7144809/). Finally, serological methods have produced a large stir across the globe with their promise of speed testing, but as of April 8^th^, 2020, WHO recommends using these tests only for research purposes (https://www.who.int/news-room/commentaries/detail/advice-on-the-use-of-point-of-care-immunodiagnostic-tests-for-covid-19), as their low sensitivity has been recalled (https://www.sciencedirect.com/science/article/abs/pii/S0033350620301141). Nonetheless, a combined genetic and serological screening is necessary to obtain a complete picture of the pandemic evolution: genetic testing to determine infected people, regardless of their symptoms; and serological testing to address immunity development (https://www.jwatch.org/na51255/2020/03/31/serologic-tests-sars-cov-2-first-steps-long-road). In the meantime, RTqPCR remains the gold standard for SARS-CoV-2 testing.

The bottleneck for SARS-CoV-2 detection by RT-qPCR is the traditional RNA isolation step (https://www.the-scientist.com/news-opinion/rna-extraction-kits-for-covid-19-tests-are-in-short-supply-in-us-67250), and here we show that it can be bypassed by performing direct 2-propanol precipitation. We have used NPSs, which has been the most common type of sample to test infection in the upper respiratory tract, but efficient detection in saliva is also documented (https://www.nature.com/articles/s41368-020-0080-z). Viruses can also be detected by an even faster method by collecting the sample with a dry swab and boiling it in lysis buffer in a closed tube, followed by direct one-step RT-qPCR using the more sensitive digital PCR (https://www.ncbi.nlm.nih.gov/pubmed/28827685). Nonetheless, this method needs validation for SARS-CoV-2 testing and care should be exercised to ensure that the lysis buffer does not interfere with the subsequent RT and PCR reactions.

RT-qPCR methods can be used to target several SARS-CoV-2 genes (https://www.ncbi.nlm.nih.gov/pmc/articles/PMC7144809/). The approach of the CDC, a US governmental agency, has focused on detecting the N gene, which encodes for the nucleocapsid, a common protein in all coronaviruses (https://www.ncbi.nlm.nih.gov/pmc/articles/PMC4147684/) using hydrolysis probes. In contrast, the approach developed by researchers at Charité Hospital (Germany), is based on detecting not only *N*, but also the RNA-dependent RNA polymerase (*RdRP*) and the envelope (*E*) genes using hydrolysis probes (https://www.ncbi.nlm.nih.gov/pubmed/31992387). In another approach recently published, researchers from the Institute for Basic Science (Korea) used intercalating dye probes to detect *N*, *E*, *RdRP* and *S*, the spike protein (https://www.ncbi.nlm.nih.gov/pubmed/3215610; we have disregarded the use of the *E* primers because of low specificity). In fact, the current Basque Government Health Surveillance protocol (April 12^th^, 2020), uses Hoffman-La Roche proprietary probes to detect *E* as a screening gene and *S* or *RdRP* as specific SARS-CoV-2 detection genes. Importantly, we have observed a large agreement of results when comparing N1 and N2 CDC’s hydrolysis probes compared to N, S, and *RdRP* Won’s intercalating dye probes. These results are very encouraging, as they suggest that different genotyping approaches used by diagnostic and research labs worldwide are in fact comparable.

Another gene that is commonly used in RT-qPCR tests is the human RNAseP subunit 30 gene *(RPP30)*. Importantly, both the CDC and Won’s probes to detect *RPP30* hybridize genomic DNA, as they do not span exon-exon junctions. In our hands, NanoDrop provided a good measurement of the quality of nucleic acids present in both the Trizol and 2-propanol extraction methods. However, a more specific detection of RNA using a Qubit 4 Fluorometer RNA integrity and quantity tests (RNA IQ assay, #Q33221; and RNA HS assay, Q32852, respectively; Invitrogen) did not show any detectable RNA in the range 250 pg/μl to 100 ng/μl (data not shown). Therefore, it is likely that in the conventional NPS preparation human RNA is largely absent or degraded, supporting the use of human genomic DNA as a sample quality control.

In conclusion, the experiments that this multidisciplinary group of researchers from the Basque Country have performed provides a simple, fast and inexpensive method to detect with high specificity and sensitivity patients infected by SARS-CoV-2. The obtained protocol shows that <u>SARS-CoV-2 positive and negative cases can be detected with precision after a simple step for precipitation of RNA, followed by one-step RT-qPCR using the CDC hydrolysis probes N1 and N2.</u>

## Data Availability

Raw data will be available upon request. The protocol is open and can be found at: https://www.ehu.eus/es/web/umayor/covid-19-protocol-en

## SUPPLEMENTARY MATERIAL

**Supplementary Figure 1.**
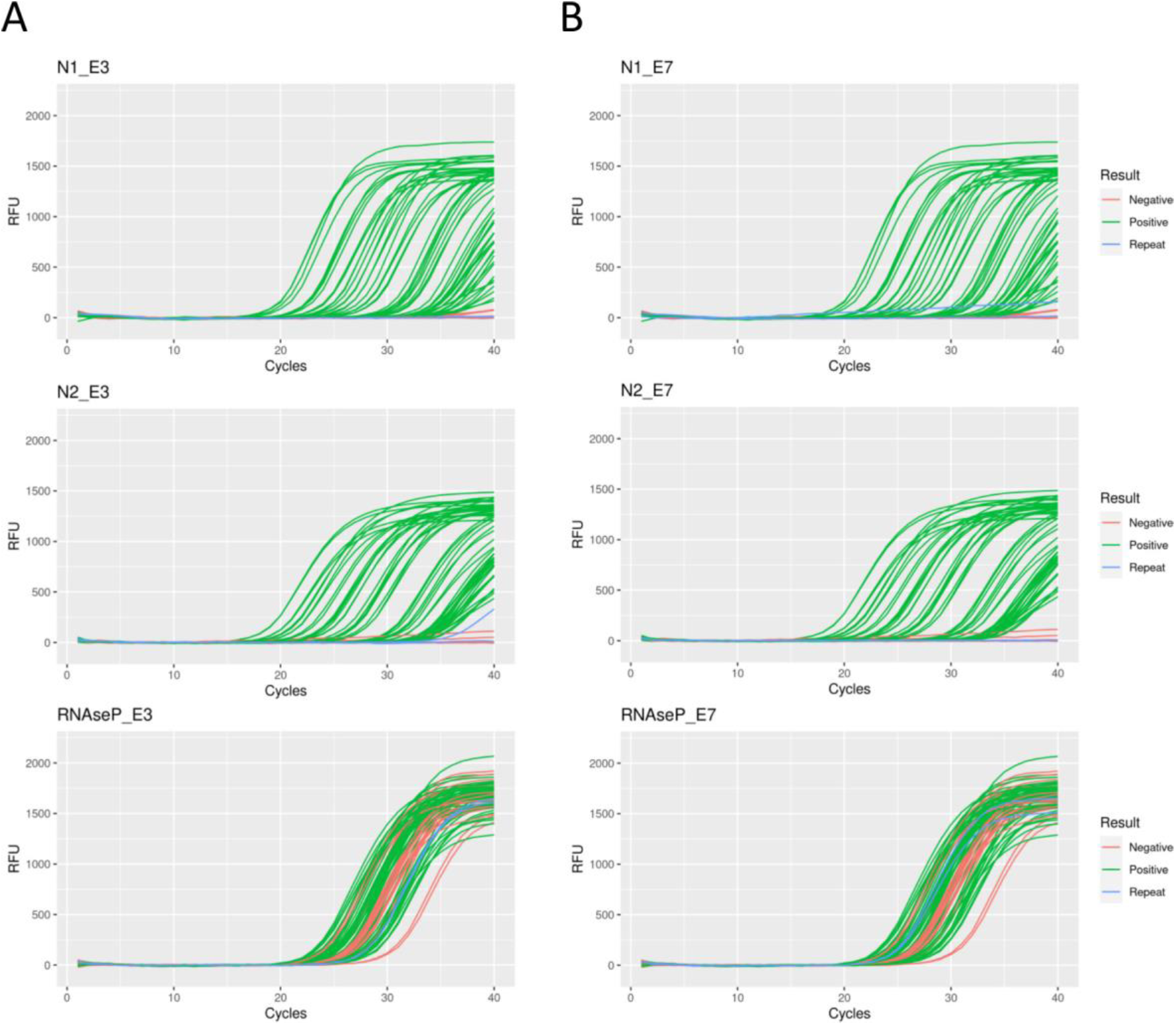
*Amplification curve analysis of hydrolysis-probe RT-qPCR results. Red, green and blue lines represent samples (in duplicate) considered negative, positive and indeterminate, respectively after the analysis with the worksheet template. In (A), the blue sigmoidal curve suggests specific but late amplification of N1 in one of the duplicates of this particular sample, and thus it should be re-analyzed with the intercalating dye method. In (B), although the blue line representing one of the replicates surpasses the detection threshold of the RT-qPCR system, it does not fit a sigmoidal curve and therefore, the sample is considered negative, without the need of re-analysis. N2 does not amplify for any of the samples whereas RNAseP (RPP30) consistently amplifies before Ct 35*.

**Supplementary Figure 2.**
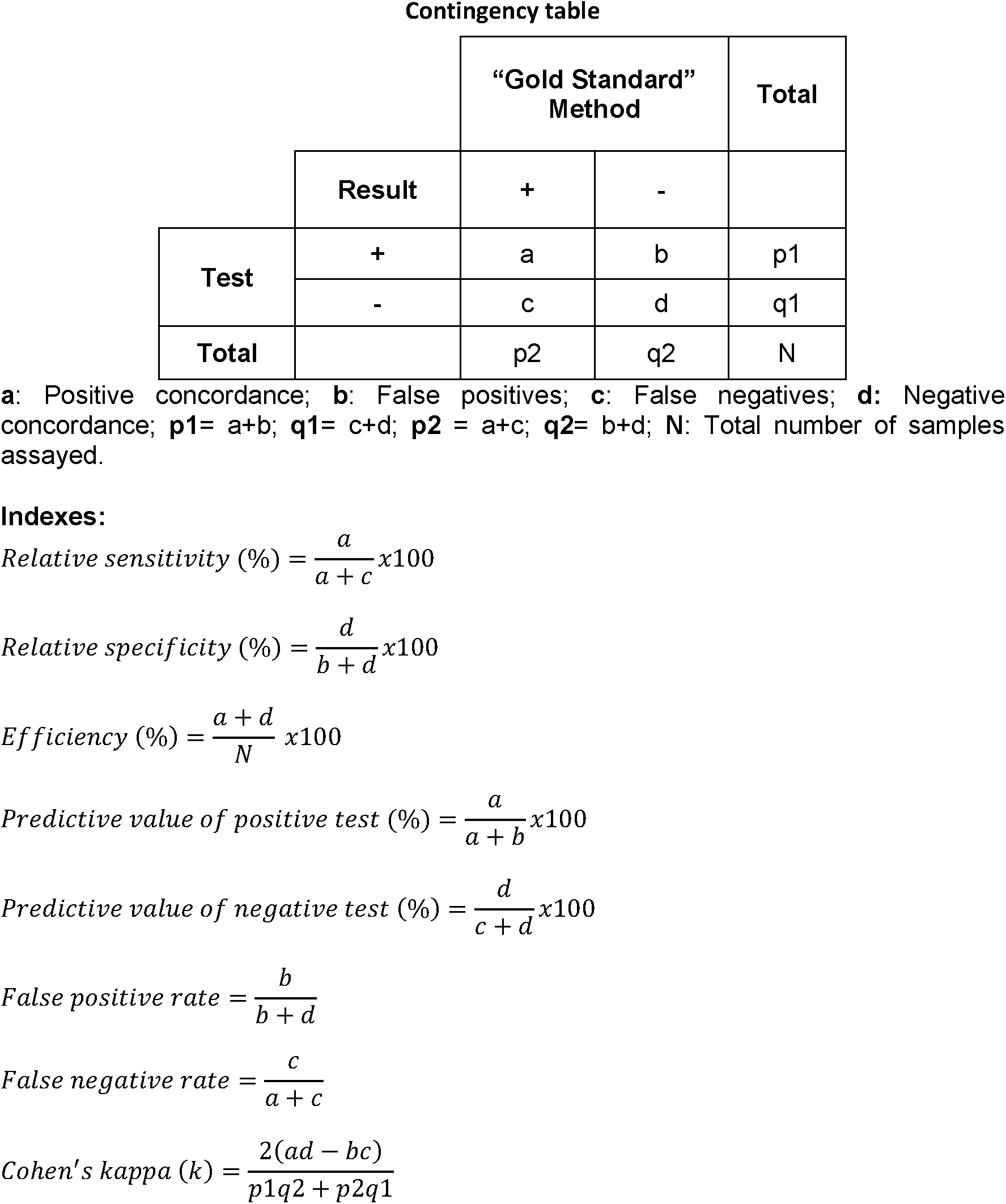
*Formulas used to calculate the different indexes of the statistical analysis*.

**Supplementary File 1.** *Excel worksheet template for semi-automated analysis of the hydrolysis probe-based experiments and assignment of clinical samples into SARS-CoV-2 positive, negative or indeterminate categories. In order for the template to function correctly, experimental plates must be prepared according to the layout described in the Raw worksheet. Sample IDs and Ct results have to be manually copied from the results file generated by the real time PCR machine and pasted into the corresponding columns*.

**Supplementary File 2.** *R script to plot qPCR curves straight from RFU raw data. To run this script RFU raw data for N1, N2 and RPP30 assays have to be exported from the qPCR system and saved in the directory where curves will be stored. Additionally, ID_well and ID_result tabs of the worksheet template provided as Supplementary File 1 will also be needed in .csv format. These .csv files should only contain information regarding the processed clinical samples (exclude controls and empty wells). All instructions are given along the script*.

## ACKNOWLEDGEMENTS AND FUNDING

This project was supported by funding from the UPV/EHU (Acción Especial “Desarrollo e implementatión del test de diagnóstico para COVID-19”). We are very grateful for the support of Nekane Balluerka, Rector of the University of the Basque Country EHU/UPV, and particularly for the support provided by José Luis Martín González (Vice-rector of Research). We are also grateful to Maitane Arantzamendi at the Microbiology Service of Cruces University Hospital (HUC, Barakaldo, Spain), Raquel Coya at the Basque BioBank and to Osakidetza for providing the inactivated sample remnants on which this protocol has been developed and validated. We are also grateful to Carlos Matute (Scientific Director) and Jaime Sagarduy (General Manager), Achucarro Basque Center for Neuroscience; and Iban Ubarretxena (Scientific Director), Biofisika Institute. Without their support this study would have not been possible.

## Notes

### Competing Interest Statement

The authors have declared no competing interest.

### Funding Statement

This project was supported by funding from the UPV/EHU (Acción Especial “Desarrollo e implementación del test de diagnóstico para COVID-19”). None of the authors received payments or services from a third party for any aspect of the submitted work.

